# Multi-Pathogen Wastewater Surveillance enables Real-Time Targeted Public Health Interventions During the 2025 African Nations Championship Football Tournament

**DOI:** 10.64898/2026.06.05.26354973

**Authors:** Andrew Nsawotebba, Innocent Morunyanga, Valeria Nakintu, Jonathan Kabazzi, Jordan Magala, Vallence Uragiwenimana, Steven Ssekyondwa, Ronald Kasujja, Harris Onywera, Noah Hull, Denis Smith Akejo, Catherine Dambya, Sulaiman Ikoba, Vito Baraka, Yenew Kebede Tebeje, Emily Barigye, Fatim Cham, Isaac Ssewanyana, Herbert Nabaasa, Allan Muruta, Charles Olaro, Diana Atwine, Susan Nabadda, Jane Ruth Acheng

## Abstract

Mass gatherings pose significant public health risks by facilitating the spread of infectious diseases. While wastewater-based surveillance (WBS) has been widely used to monitor pathogens in high-income settings, its use as a practical, multi-pathogen surveillance tool during mass gatherings in low- and middle-income countries remains limited. This study aimed to assess the operational feasibility, epidemiological significance, and public health utility of multi-pathogen WBS during the African Nations Championship (CHAN) football tournament in Uganda. Wastewater surveillance was conducted at Mandela National Stadium during eight match days in August 2025. Moore swabs were deployed at 38 manholes receiving wastewater from different toilet facilities across the stadium to capture representative wastewater samples. Samples were processed using Nanotrap® microbiome virus particles to concentrate pathogens, followed by nucleic acid extraction. Samples were analyzed for multiple enteric and respiratory pathogens, including Mpox, using quantitative PCR (qPCR). Descriptive analyses were performed to characterize pathogen detection patterns, positivity rates, and temporal distribution across surveillance sites. A total of 304 wastewater samples were collected and analyzed, of which 259 (85.2%) tested positive for at least one pathogen. Multiple pathogens were consistently detected across sampling days, with enteric pathogens predominating, particularly *Shigella* spp. (53.6%), Rotavirus A (35.9%) and Enterovirus (32.2%). The mpox virus was also detected in a notable proportion of samples (28.6%) across several sampling days. Respiratory pathogens, including SARS-CoV-2 (11.8%) and Influenza B (8.2%), were identified intermittently at lower frequencies. Pathogen diversity varied over time, with up to eight pathogens detected on a single day, and co-detection of multiple pathogens observed in the majority of positive samples. Cq value distributions further demonstrated variability in detected signal patterns across pathogens. Surveillance findings informed real-time public health interventions, including sanitation reinforcement, intensified hygiene promotion, environmental disinfection, and targeted risk communication, strengthened syndromic surveillance with on-site triage, and targeted environmental health assessments of food handling and wastewater infrastructure. These findings demonstrate the operational feasibility and public health utility of integrating multi-pathogen wastewater-based surveillance into mass-gathering preparedness and response frameworks in low-resource settings. By capturing diverse pathogen signals and informing targeted interventions during the CHAN football tournament, WBS can provide actionable population-level insights that can support outbreak preparedness and response. Scaling WBS within national preparedness systems could strengthen epidemic intelligence, enhance early warning capacity, and support data-driven public health decision-making during future mass gatherings and emerging infectious disease threats.

## Background

Wastewater-based surveillance (WBS), which involves monitoring pathogens in sewage or other human-wastewater-impacted environmental waters, has emerged as a critical component of public health surveillance systems [1]. WBS is not a new field; however, the benefits of WBS for infectious disease surveillance became more widely recognized during the COVID-19 pandemic [2]. WBS has been used to monitor water-borne or fecal-orally transmitted pathogens, offering a sensitive way to observe changes and varieties of pathogens within communities, such as poliovirus [3]. Regular large-scale screening in a clinical setting poses challenges, and individuals who are asymptomatic or have mild symptoms often remain undetected [4]. WBS captures shedding from both symptomatic and asymptomatic individuals, is cost-effective, scalable, and provides representative population-level data when samples are collected from strategic sites [5,6]. In addition, analyzing sewage samples does not require informed consent, thereby limiting ethical concerns and reducing practical and logistical barriers to sampling [7].

The success of WBS in recent years has raised concerns about its potential extension to other infectious agents and contexts, including the public health surveillance of mass gathering events [8]. Mass gatherings (MGs) are events attended by a large number of people, which can pose public health risks and strain public health resources in the host country [9]. Such events typically bring together dense populations with frequent close contact and significant movement of participants from both local and international settings, creating conditions that favor the rapid spread of infectious diseases [10]. MGs occur in various contexts, encompassing sporting, cultural, religious, and political domains, and are recognized for exerting considerable strain on the host country’s economy, infrastructure, and health system [11]. Previous MGs have demonstrated the value of WBS as a complementary tool for outbreak preparedness and response. For instance, WBS during the FIFA World Cup Qatar 2022 highlighted the need for enhanced surveillance of infectious diseases among large international crowds [12]. Despite successful applications in high-income settings, evidence from low- and middle-income countries (LMICs) remains scarce. This study addresses this gap by implementing multi-pathogen WBS during the African Nations Championship Football Tournament.

The African Nations Championship (CHAN) is a biennial football tournament organized by the Confederation of African Football (CAF). It was announced in 2007 and inaugurated in 2009. Unlike the Africa Cup of Nations, participation in CHAN is restricted to players active in their home country’s domestic leagues, making it distinct in its representation of local football talent [13]. The tournament attracted many spectators and participants, creating crowded gatherings that can heighten public health concerns. Uganda, along with Kenya and Tanzania, co-hosted the CHAN tournament in August 2025. Matches in Uganda were held at the Mandela National Stadium in Namboole, Kampala, drawing large numbers of players, officials, spectators, and visitors from across the country and across Africa. These crowds extended to accommodations, transport hubs, and nearby communities, increasing the risk of the spread of infectious diseases. To enhance public health readiness and preparedness during and after the tournament, the Ministry of Health’s Departments of National Health Laboratory & Diagnostic Services (NHLDS) and Environmental Health jointly conducted a WBS in August 2025. This study aimed to demonstrate the operational feasibility, epidemiological significance, and public health utility of multi-pathogen WBS during the CHAN football tournament in Uganda.

## Materials and Methods

### Study Design and Setting

WBS was conducted during the CHAN football tournament, held August 4–29, 2025, at Mandela National Stadium in Namboole, Kampala, Uganda. The wastewater infrastructure is designed to collect sewage from multiple toilet facilities distributed throughout the stadium. The facility comprises 38 toilets located in different sections, each draining into a dedicated manhole as shown in Fig 1. These tributary manholes converge into a single main manhole, which aggregates wastewater from all stadium facilities before discharging it into a nearby lagoon for initial containment and subsequent treatment or release. Wastewater sampling was conducted on the eight Uganda match days of the tournament, which were selected to coincide with periods of maximum stadium occupancy and peak wastewater generation. Sampling was conducted on the following match days: 4th August, 8th August, 11th August, 15th August, 18th August, 23rd August, 26th August, and 29th August, 2025. Non-match days were not sampled because the stadium was not in active use for tournament events on those days. This strategy prioritized capturing pathogen signals at maximum population density rather than achieving continuous temporal coverage throughout the tournament period.

**Fig 1:**
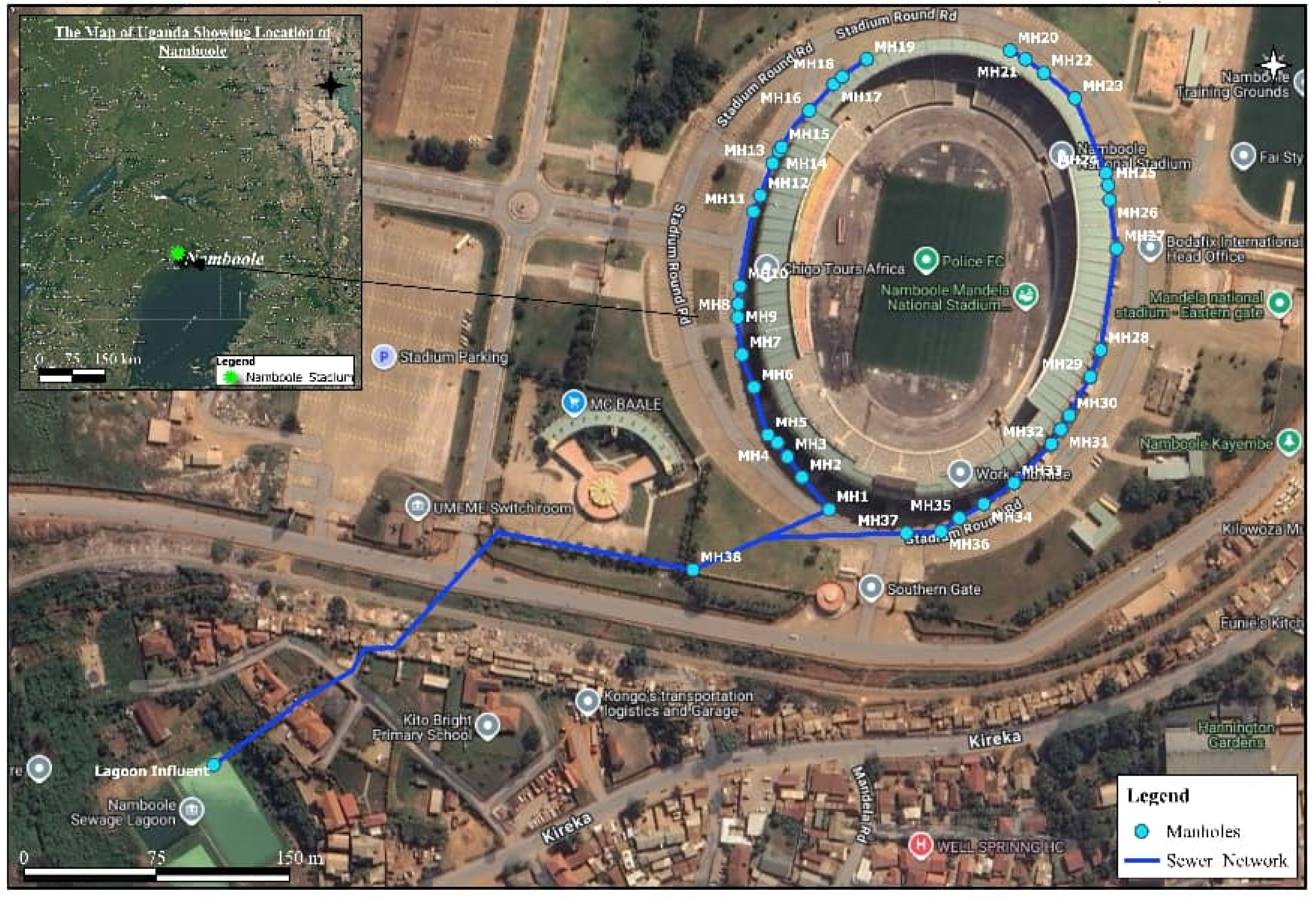
Schematic diagram of the wastewater management system and sampling points at Mandela National Stadium, Namboole, Kampala, Uganda. Numbered markers indicate the 38 manholes sampled during the CHAN 2025 football tournament. The main aggregation manhole (arrowed) collects combined flow from all tributary manholes before discharge to the lagoon.

This wastewater flow configuration informed the sampling strategy. A cross-sectional study design was employed, and 38 manholes were selected for sampling because they represent the primary collection points for wastewater from individual toilet facilities. By sampling all major tributary manholes, the strategy ensured comprehensive capture of wastewater from all functional toilet facilities across the stadium, thereby providing representative samples that reflected the stadium population at the time of sampling. No formal a priori sample size calculation was performed. All 38 manholes were successfully sampled on each of the eight match days, yielding 304 samples with no missing observations.

### Justification of the pathogen diagnostic Panel

Public health priorities for MGs in Uganda informed the selection of this specific pathogen panel. All targeted pathogens are causative agents of priority notifiable and epidemic-prone diseases under Uganda’s Integrated Disease Surveillance and Response (IDSR) system [14]. Moreover, their inclusion was further justified by their high potential to rapidly spread in crowded settings, given their known transmission routes, including fecal-oral, respiratory, and direct contact, all of which are well-documented risks in MGs [15].

### Sample collection

Moore swab sampling was employed as the wastewater collection method. Sampling was conducted throughout the tournament at the stadium to maximize the likelihood of detecting pathogens introduced or amplified by the mass-gathering population. Each Moore swab was folded into an 8-ply pad, secured with a string long enough to reach the wastewater influent, autoclaved, and sealed in a Ziploc® bag [16]. At the sampling sites, the swab was tied to a stable structure and fully submerged in the wastewater for 24 hours. Installations and retrievals were carried out between 7:00 a.m. and 9:00 a.m. After the 24-hour exposure period, the swab was placed in a clean Ziploc® bag for transport [17]. All external surfaces were disinfected with 70% ethanol before placing the samples into bio-bottles and transporting them in an ice-cold (2-8 °C) box to the Central Emergency Response and Surveillance Laboratory (CERSL), located approximately 16 km from the sampling sites. Moore swab sampling was conducted once per site per day.

### Sample Processing

Specimen processing and testing were performed at CERSL within 1 hour of collection. Upon receipt, the absorbed wastewater was aseptically expressed from each Moore swab without removing it from its sterile Ziploc® bag. The bag was aseptically opened, and the contents were transferred to a sterile 500 mL flask. External pressure was applied to compress the swab and release the liquid, yielding approximately 250 mL per sample. The expressed liquid from the Moore swabs was then concentrated.

### Nucleic acid concentration, capture, and extraction/purification

From each collected wastewater sample, 35 mL was aliquoted into a 50 mL tube for concentration using a magnetic bead–based method. Briefly, 100 µL of Nanotrap® Enhancement Reagent 2 (ER2) (SKU# 10112, Ceres Nanoscience, Inc., Manassas, VA) was added to the aliquot and vortexed. Subsequently, 525 µL of Nanotrap Microbiome Particles (SKU#44202, Ceres Nanosciences, Inc., Manassas, VA, USA) was added; the mixture was inverted to ensure a uniform suspension and then incubated at room temperature for 10 minutes. Viral particles bound to the beads were pelleted using a DynaMag™-50 magnet, then eluted in 1 mL of Nanotrap Buffer 2. The eluted viral particles were lysed and purified using the Qiagen Viral DNA Mini kit or the Qiagen QIAamp Viral RNA Mini kit (Qiagen, Hilden, Germany), as applicable, following the manufacturer’s instructions. Nucleic acids were eluted with the manufacturer-provided AVE buffer to a final volume of 60 µL. A negative extraction control was included during the extraction process. Pepper mild mottle virus (PMMoV) was used as an internal process control to monitor nucleic acid extraction efficiency and assess PCR inhibition across all samples.

### PCR Assays, Cycling Conditions, and Results Analysis

The extracted nucleic acids were amplified on a Bio-Rad CFX 96 Dx real-time thermal cycler (Bio-Rad Laboratories, Hercules, CA, USA) using detection kits. To detect target pathogens, singleplex and multiplex real-time PCR kits from Jiangsu Bioperfectus Technologies (Jiangsu Province, China) were used. Master mixes were prepared in a nuclease-free environment according to the manufacturer’s instructions. PCR amplification conditions were set according to the manufacturer’s recommendations for each kit. To ensure analytical validity, each run included non-template controls (NTCs) and manufacturer-provided negative and positive controls. A sample was considered positive for a target pathogen if it produced a characteristic amplification curve with a quantification cycle (Cq) value at or below the manufacturer-specified cutoff for the respective kit. Because the aim was qualitative nucleic acid detection rather than quantification, we used the manufacturer’s established limit of detection (LoD) for all assays.

**Table 1:**
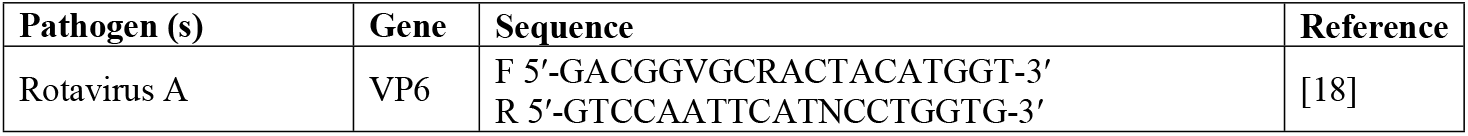
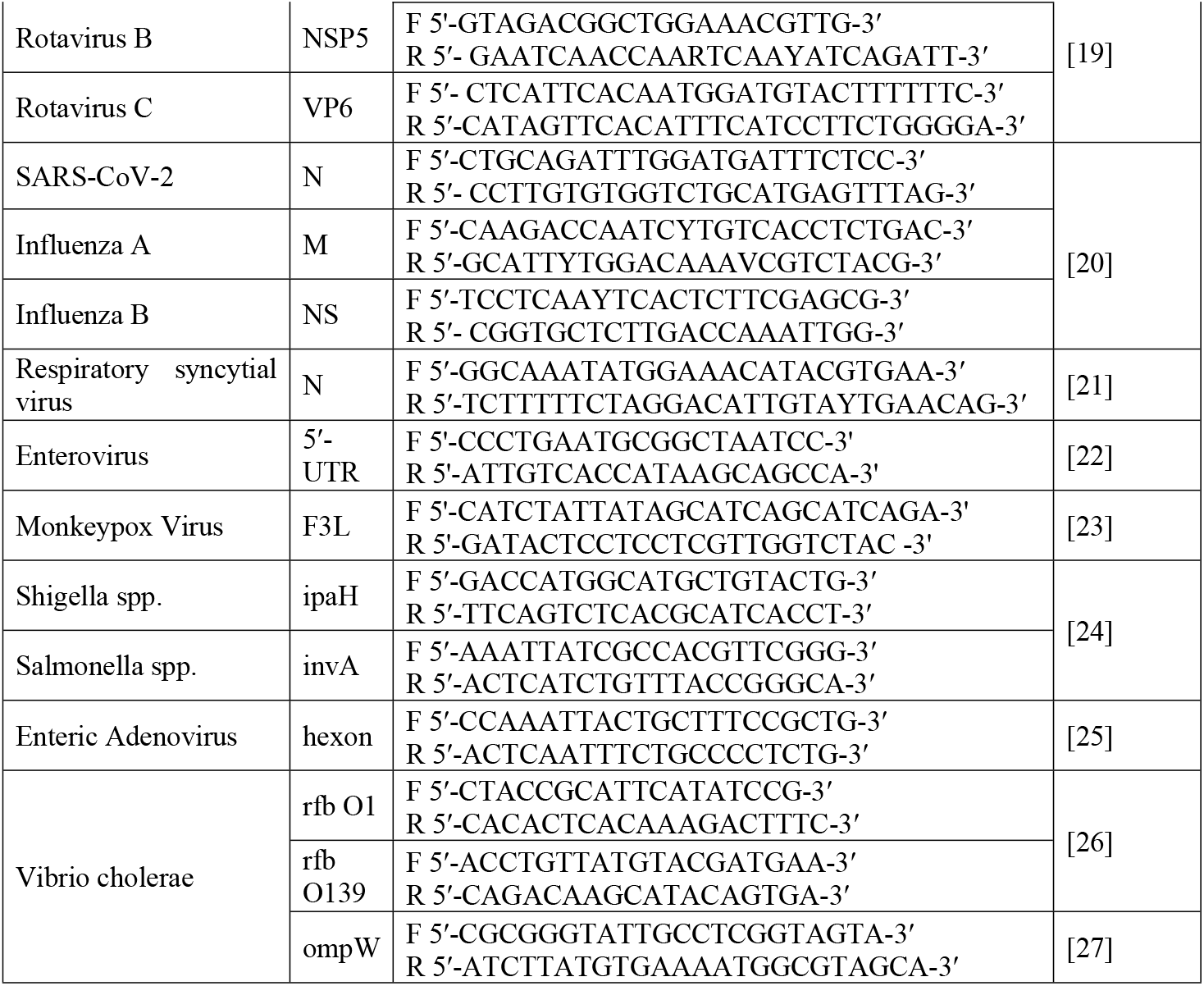
PCR primer sequences for pathogen detection assays.

### Data Analysis and Visualization

Data analysis and visualization were performed using R statistical software (R Foundation for Statistical Computing, Vienna, Austria; version 4.5.1, 2025), with visualizations generated using the *ggplot2* package. RStudio (version 2025.09.1+401) was used as the integrated development environment. Descriptive statistical analyses were performed to summarize pathogen detection frequencies and proportions across sampling days and sites. Pathogen positivity was calculated as the proportion of samples testing positive for each target pathogen. Co-detection patterns were evaluated based on the number of pathogens detected per sample. Data visualization included stacked bar charts to illustrate daily pathogen distribution and horizontal bar charts to present overall pathogen positivity. Given the descriptive and operational nature of this study, formal inferential statistical analyses were not conducted. All 304 samples that met quality control criteria were included in the analysis; no samples were excluded due to extraction or amplification failure. No subgroup, interaction, or sensitivity analyses were conducted, consistent with the descriptive study objective.

## Results

A total of 304 wastewater samples were collected across all manholes at Mandela National Stadium during the surveillance period, of which 259 (85.2%) tested positive for at least one pathogen.

### Proportions of positive pathogens detected daily

The stacked bar chart (Fig 2) illustrates the daily distribution and proportional contribution of each pathogen over eight consecutive sampling days. Enteric pathogens, particularly Shigella spp. and Rotavirus A, consistently contributed a substantial proportion of detections across multiple days. Respiratory pathogens, including SARS-CoV-2 and Influenza B, were detected intermittently at lower frequencies. The highest pathogen detection was recorded on August 23, 2025, when eight distinct pathogens were identified, which coincided with peak attendance. In contrast, only Enterovirus was detected on August 26, 2025. The data highlight considerable variation in pathogen diversity detected during the surveillance period.

**Fig 2:**
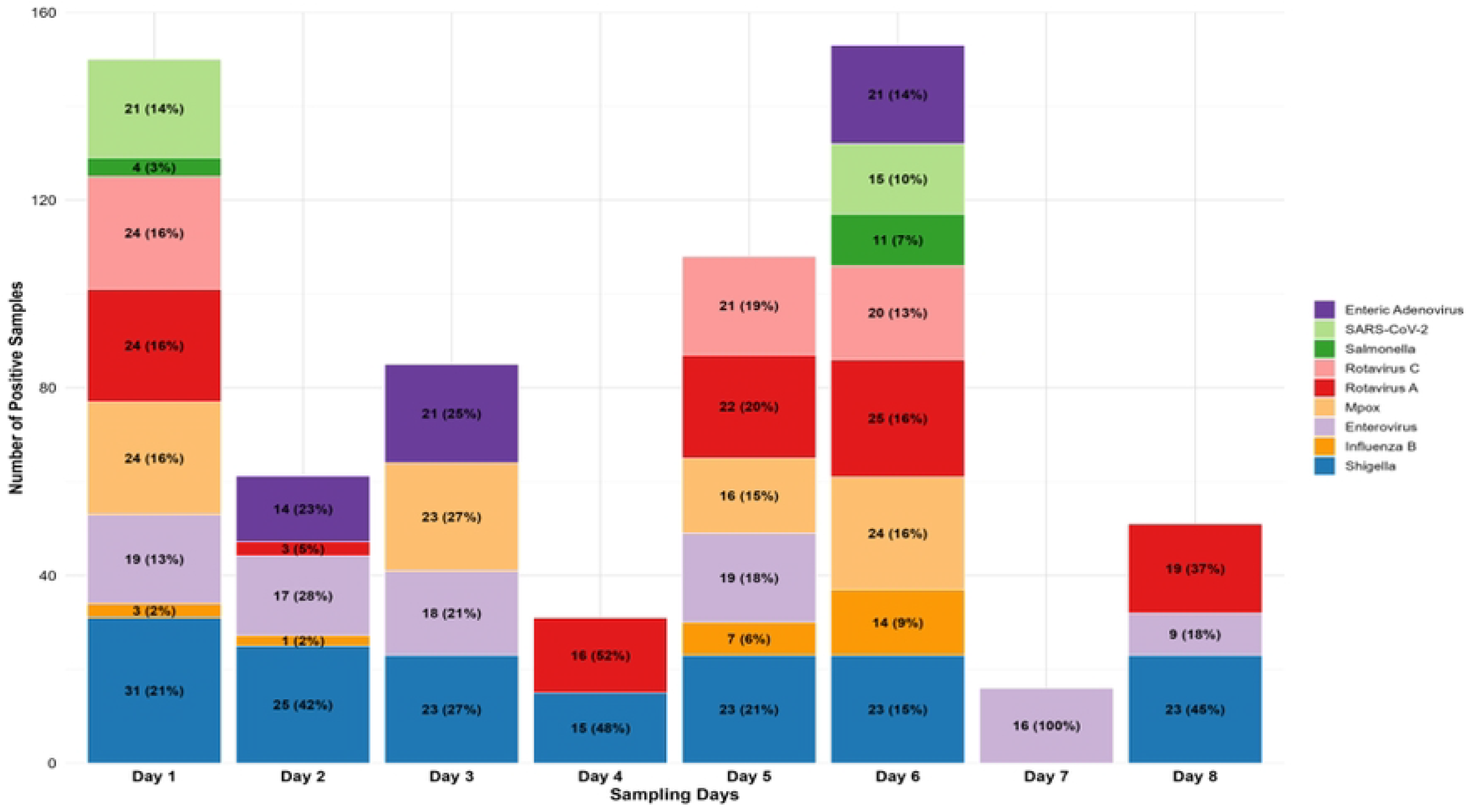
Stacked bar chart showing the daily distribution and proportional contribution of each detected pathogen. The y-axis represents the total number of positive detections per day across all 38 manholes. Each color segment represents a distinct pathogen target. Pathogens not detected on a given day are absent from the corresponding bar.

### Pathogen burden and average positivity percentage across all sites

The analysis of aggregated pathogen counts and average positivity percentages across all sites showed that enteric bacterial pathogens were the most common, with Shigella spp. detected most frequently (n = 163, 53.6%). (Fig 3). Among viral pathogens, Rotavirus A (n = 109, 35.9%) and Enterovirus (n = 98, 32.2%) were most prevalent, while Rotavirus C was detected at a lower frequency (n = 65, 21.4%). Enteric Adenovirus was also detected (n = 56, 18.4%), although at a lower frequency. Notably, Mpox was also detected in a considerable proportion of samples (n = 87, 28.6%), suggesting circulation within the contributing population during the mass gathering. Other bacterial pathogens appeared less often, with *Salmonella* spp. detected at (n = 15, 4.9%). Respiratory pathogens were detected at lower frequencies, with SARS-CoV-2 identified in 36 cases (11.8%) and Influenza B in 25 cases (8.2%), consistent with intermittent detection during the surveillance period. Rotavirus B, Influenza A, Respiratory Syncytial Virus (RSV), and *Vibrio cholerae* were not detected in any of the samples during the surveillance period.

**Fig 3:**
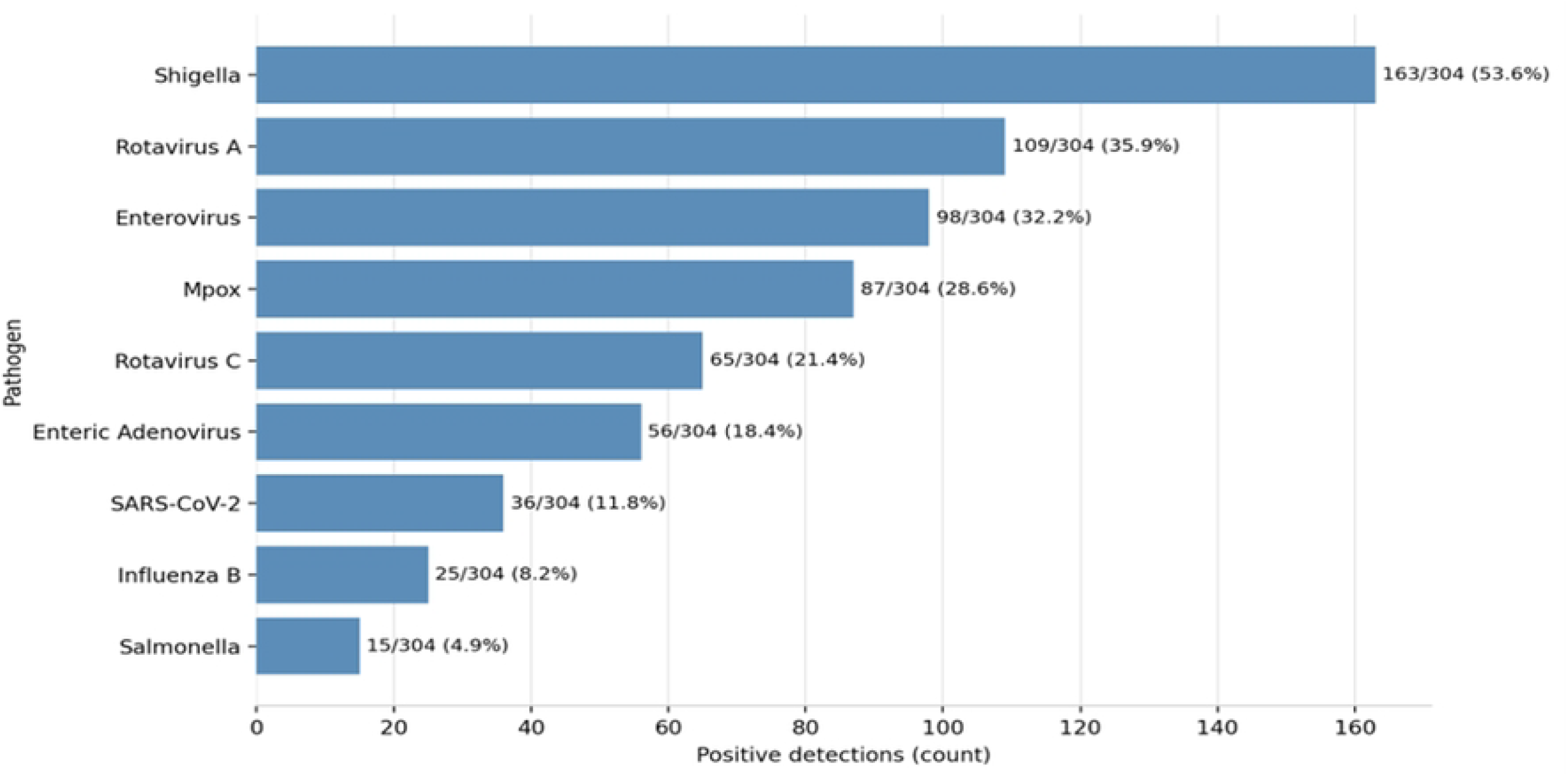
Horizontal bar chart showing the frequency (total positive sample count, top axis) and average positivity percentage (bottom axis) for each detected pathogen (n=304 samples). Pathogens are ordered from highest to lowest overall positivity. Pathogens not detected in any sample (Rotavirus B, Influenza A, RSV, Vibrio cholerae) are not shown.

### Cq value distributions

Among pathogens with at least one positive detection, Cq values varied, reflecting differences in target nucleic acid concentrations within the wastewater matrix (Fig 4).

**Fig 4:**
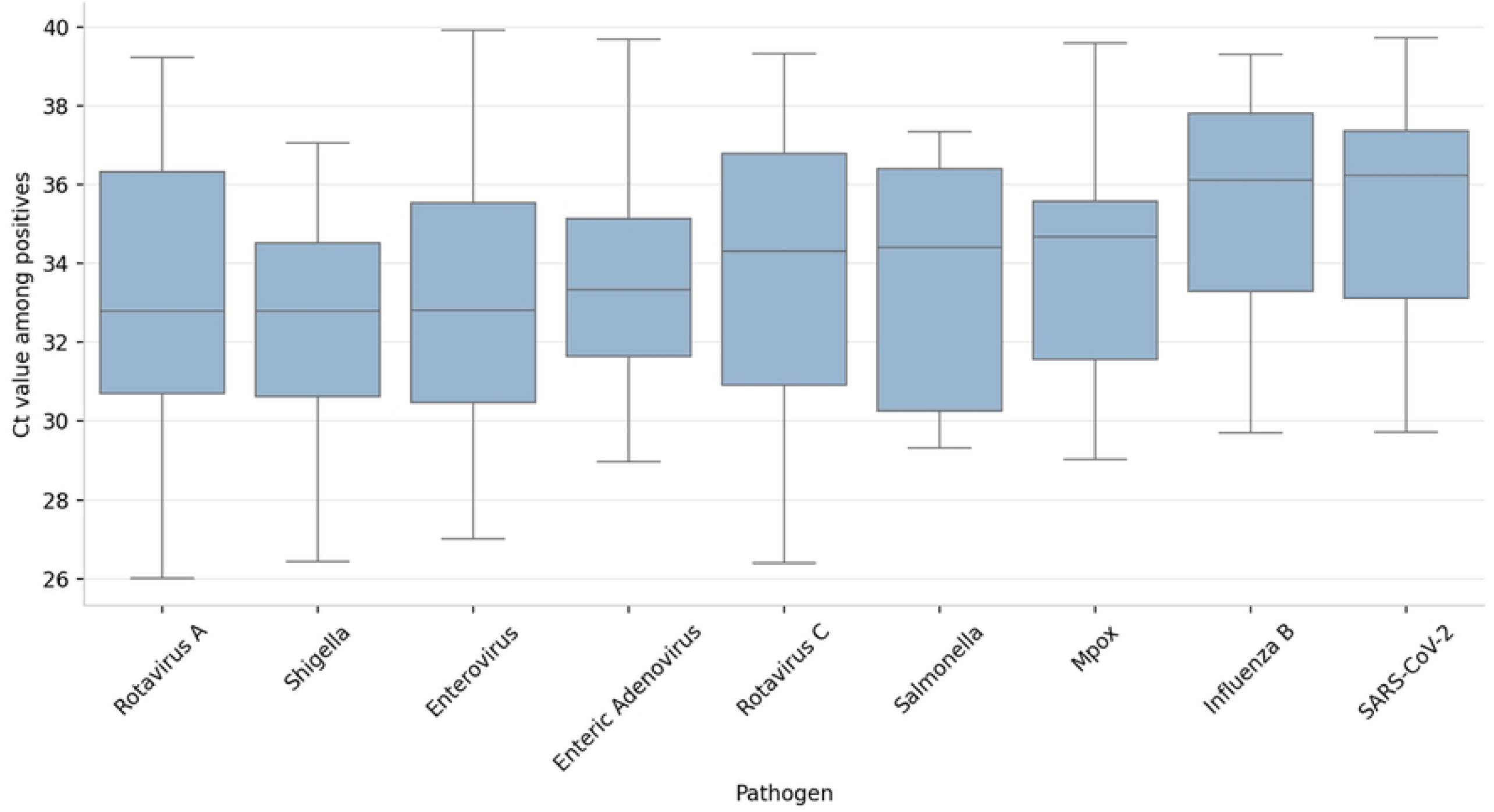
Box-and-whisker plots showing the distribution of Cq values among positive detections for each pathogen. The horizontal line within each box represents the median Cq value; box limits represent the interquartile range (IQR); whiskers extend to 1.5 × IQR; individual outliers are plotted as points. Higher Cq values indicate lower nucleic acid concentrations in the wastewater matrix.

Rotavirus A and Shigella spp. exhibited the lowest median Cq values (both 32.8; IQR 30.7–36.3 and 30.6–34.5, respectively), with Enterovirus (median Cq = 33.0; IQR 30.5–35.6), Enteric Adenovirus (median Cq = 33.3; IQR 31.6–35.1), and Rotavirus C (median Cq = 34.3; IQR 30.9– 36.8) showing similar Cq distributions. In contrast, higher median Cq values were observed for SARS-CoV-2 (median 36.4; IQR 33.4–37.5) and Influenza B (median 36.1; IQR 33.3–37.8). The mpox virus demonstrated an intermediate Cq distribution (median 34.7; IQR 31.6–35.6). Overall, Cq values showed variability within and between pathogen groups.

### Co-detection of multiple pathogens

The distribution of the number of pathogens detected per sample is shown in Fig 5. Among positive samples, multiple pathogens were frequently detected within the same sample. A total of 80 samples had a single pathogen detected, while the majority showed co-detection of two or more pathogens. Specifically, 66 samples had two pathogens, 46 had three, 38 had four, 22 had five, and 7 samples had six pathogens detected.

**Fig 5:**
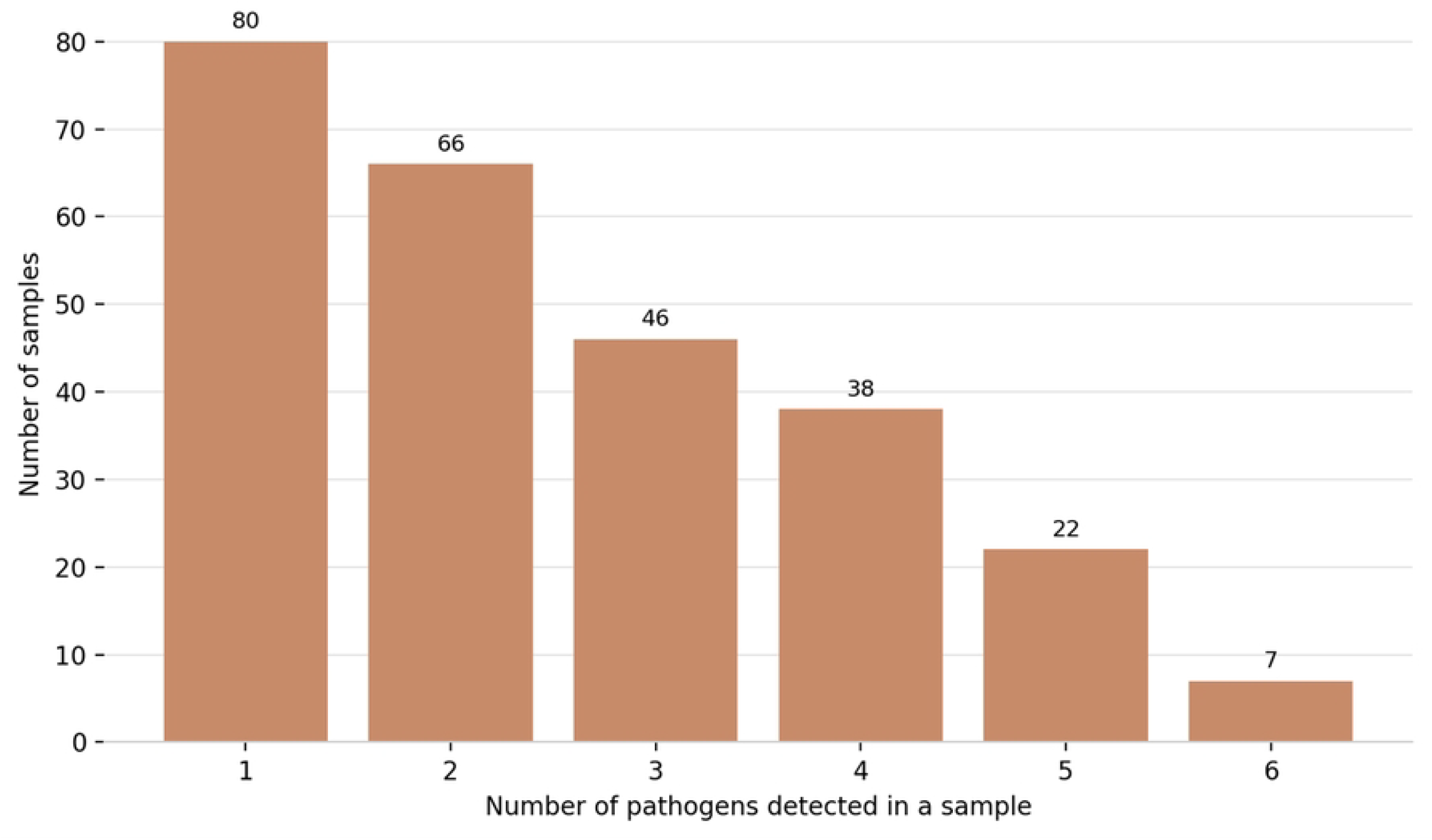
Bar chart showing the distribution of the number of pathogens co-detected within individual positive wastewater samples (n=259 positive samples). The x-axis indicates the number of pathogens detected per sample; the y-axis shows the count of samples in each category. The proportion of positive samples co-detected with two or more pathogens (69.1%, 179/259) is annotated.

Overall, 179 out of 259 positive samples (69.1%) exhibited co-detection of at least two pathogens, indicating that multi-pathogen presence within individual wastewater samples was common during the surveillance period.

## Discussion

The CHAN football tournament held in Uganda in August 2025 provided a critical opportunity to assess the value of WBS as a public health tool during MGs in a sub-Saharan African setting. This study successfully captured real-time signals of enteric, respiratory, and emerging pathogens at a major sporting event, highlighting the complex microbial landscape of such settings.

The detection of nine distinct pathogens, including *Salmonella, Shigella* spp., Rotavirus A, Rotavirus C, Enterovirus, Enteric Adenovirus, mpox virus, SARS-CoV-2, and Influenza B, demonstrates the diversity of pathogens detected in wastewater during MGs. The predominance of enteric pathogens in wastewater during the surveillance period, particularly Shigella spp., is consistent with previous studies reporting the detection of enteric pathogens in wastewater [28] and highlights the potential for faecal–oral transmission in MG settings [29]. These pathogens are commonly associated with gastrointestinal infections [30]. Their detection in wastewater may indicate their presence within the contributing population during the surveillance period. In mass gathering settings, where large numbers of individuals share sanitation infrastructure, this underscores the importance of strengthened hygiene measures, food safety protocols, and environmental health monitoring.

The detection of the mpox virus in wastewater across multiple sampling days indicates its presence within the contributing population during the surveillance period, despite a decline in reported clinical cases as of August 2025 [31]. This finding is consistent with previous studies that have identified mpox through environmental surveillance, even in the context of limited clinical reporting [32]. The high spatial positivity (87 of 304 samples, 28.6%) should be interpreted in the context of wastewater’s aggregated nature: a single infected individual shedding mpox virus may contribute a signal to multiple manholes depending on toilet usage patterns and wastewater flow connectivity. The proportion of positive manholes, therefore, reflects the spatial reach of mpox-positive wastewater contributions rather than a direct estimate of mpox prevalence among stadium attendees. Nonetheless, the detection of mpox signal across multiple sites and multiple sampling days — including on days not coinciding with the highest overall pathogen burden — is consistent with sustained low-level circulation within the contributing population during the tournament period, warranting the public health response described above. Quantitative genomic analysis (genome copies per litre) in future studies would enable more refined estimation of pathogen burden from wastewater signal intensity.

The detection of respiratory pathogens at low frequencies, including SARS-CoV-2 and Influenza B, demonstrates that viral respiratory infections remain significant risks in mass gatherings [11].

Co-detection of multiple pathogens was a prominent feature of this study, with the majority of positive samples containing two or more pathogens. This observation reflects the complex composition of wastewater, which integrates contributions from multiple individuals and sources [1,33]. Rather than representing individual co-infections, these findings highlight WBS’s capacity to capture overlapping pathogen signals within a shared environmental matrix, providing a more comprehensive picture of pathogen presence during mass gatherings.

The observed variability in Cq values across pathogens further supports heterogeneity in the signals detected within the wastewater system. While lower Cq values were observed for some enteric pathogens and higher values for respiratory pathogens, these differences should be interpreted cautiously, as Cq values are assay-dependent and not directly comparable across targets. Nonetheless, the Cq distributions provide useful descriptive insight into relative signal patterns and variability across pathogens during the surveillance period.

Variation in pathogen detection across sampling days was also observed, with higher levels recorded on matchdays involving Uganda, which coincided with increased attendance. While this temporal alignment suggests a potential association between crowd density and pathogen detection patterns, these observations should be interpreted with caution, given the absence of baseline data and the study’s descriptive nature.

Although WBS has been used in high-resource MG settings, these implementations have generally been limited to high-income countries, for instance, SARS-CoV-2 monitoring in Tokyo’s strictly regulated Olympic Village [34] and on SARS-CoV-2 and poliovirus monitoring during the 2022 FIFA World Cup in Qatar [12]. In contrast, this study demonstrates the feasibility and utility of a multi-pathogen WBS approach in an LMIC context, reflecting real-world public health conditions. This work builds on existing African WBS literature, such as Cape Town’s use of WBS for SARS-CoV-2 during a sports gathering, proving the adaptability of this tool across the continent’s diverse public health landscapes [35].

### Public Health Interventions Triggered by the WBS findings

During the CHAN football tournament, WBS enabled the real-time activation of targeted public health interventions. Implemented across the eight match days, these actions translated surveillance intelligence into rapid containment measures, tailored to pathogen detections and site-specific risks.

### Risk Communication and Community Engagement

The sustained detection of enteric pathogens, including Shigella spp., Rotavirus A, and Rotavirus C, and the emergence of mpox in wastewater prompted intensified risk communication and community engagement (RCCE) activities. Health messages promoting handwashing and respiratory hygiene were disseminated via stadium announcements, digital signage, and posters. Following the detection of the mpox virus, official media briefings were held to raise public awareness, promote preventive behaviors, and encourage self-monitoring for symptoms.

### Enhanced Hygiene & Sanitation Protocols

Repeated detection of enteric pathogens prompted the escalation of hygiene and sanitation measures within the stadium. Mobile handwashing stations and alcohol-based sanitizers were deployed at key locations, including toilets, food vending areas, and entrances. Toilet cleaning frequency was increased, with targeted disinfection between matches. Routine cleaning of high-touch surfaces, such as railings, seating areas, and vendor stalls, was performed using chlorine-based solutions. Preventive inspections of wastewater manholes and containment systems were also conducted by the Ministry of Health’s Environmental Department to mitigate the risk of overflow.

### Clinical Surveillance & Syndromic Linkage

The detection of multiple pathogens in wastewater was associated with strengthened syndromic surveillance. On-site triage and point-of-care testing were conducted for symptomatic individuals, with established protocols for clinical management and referral. Community engagement teams were also deployed to support symptom screening for dermatological and respiratory conditions.

### Field Investigations & Environmental Assessments

In response to the pathogen profiles identified in wastewater, targeted field investigations and environmental assessments were undertaken. Environmental health officers conducted sanitation assessments and conducted spot checks on food-handling and water-safety practices. Continuous monitoring of the wastewater system was maintained to track pathogen trends. At the same time, inter-agency coordination among the Ministry of Health, stadium management, and security teams ensured a coordinated, responsive public health approach.

### Public Health Integration

The findings of this study provide a practical pathway for integrating WBS into Uganda’s national MG preparedness frameworks. Building on existing expertise from environmental surveillance in Uganda [36,37], a structured WBS framework can be developed and operationalized for MG settings. Such a framework would include key phases: pre-event baseline sampling at priority venues to establish background pathogen levels; real-time event-based monitoring with predefined pathogen-specific alert thresholds; a formal response protocol linking WBS findings to timely public health actions; and post-event surveillance.

Beyond acute infectious disease monitoring, WBS also offers an opportunity to assess the community-level burden and emergence of antimicrobial resistance (AMR). Wastewater samples can be further analyzed using metagenomic sequencing or targeted qPCR to characterize the abundance and diversity of AMR genes circulating in populations during mass gatherings. This approach aligns with global initiatives such as the WHO Global Antimicrobial Resistance Surveillance System (GLASS), which promotes integrated, population-level surveillance strategies to strengthen AMR intelligence [38]. The findings from this study are most directly applicable to stadiums and enclosed venues with identifiable tributary wastewater infrastructure. Adaptation to settings with less-structured drainage systems — including informal settlements and open-air gathering sites will require modified sampling strategies.

### Limitations

The absence of environmental baseline data from the pre-event period limits the ability to compare pathogen signals attributable to the MG directly. This was due to logistical and financial constraints associated with the rapid deployment of surveillance in this resource-limited setting. The lack of quantitative pathogen measurements (genome copies per mL) further constrained detailed assessment of pathogen dynamics. Additionally, environmental surveillance findings were not directly linked to clinical outcomes, although the implementation of onsite syndromic surveillance provided partial contextual support. Sampling was restricted to the eight Uganda match days and did not capture potential pathogen dynamics on non-match days or during the post-event period. Continuous sampling across all tournament days would provide a more complete picture of pathogen dynamics over the tournament’s full temporal arc [33]. Future studies with pre-event baseline data and larger sample sizes will be better positioned to statistically assess temporal trends and associations in pathogen co-occurrence. Cq values were used for descriptive comparison only and are assay-dependent; therefore, they are not directly comparable across pathogens and do not represent quantitative measures of pathogen concentration. Additionally, the detection of multiple pathogens within a single wastewater sample reflects the composite nature of wastewater and does not represent individual-level co-infection. Moore swab capture efficiency may vary with wastewater flow rate and organic load, introducing variability in pathogen detection across sampling sites and days. Environmental conditions (temperature, rainfall, wastewater flow rate) during sampling days were not systematically recorded. These factors are known to influence Moore swab capture efficiency and the stability of nucleic acids in wastewater matrices. They may have contributed to some of the observed day-to-day variability in pathogen detection. Despite these limitations, the successful implementation of a broad, multi-pathogen surveillance approach in a typical, open-city LMIC context demonstrates that WBS is operationally feasible and highly informative during MGs.

## Conclusion

This study demonstrates that multi-pathogen wastewater-based surveillance can be effectively implemented in a real-world MG setting in a low- and middle-income country, providing actionable public health intelligence in near-real time. The high overall positivity rate (85.2%) and detection of nine distinct pathogens highlight the intensity and diversity of pathogen circulation within the stadium environment during the CHAN football tournament. Importantly, integrating WBS findings with rapid public health responses demonstrates the operational value of this approach beyond passive monitoring.

These findings position WBS as a practical and scalable tool for mass gathering preparedness and response, particularly in resource-limited settings where routine clinical surveillance may be constrained. Future efforts should focus on integrating WBS into national surveillance frameworks, establishing baseline datasets, and incorporating quantitative and genomic approaches to enhance resolution and early warning capacity. Expanding this model across diverse mass gathering contexts will be critical to strengthening epidemic preparedness and supporting data-driven public health decision-making.

## Acknowledgements

We acknowledge the Ministry of Health, the Department of National Health Laboratory and Diagnostic Services, and the Environmental Health for coordinating the wastewater surveillance activities during the African Nations Championship football tournament. We gratefully acknowledge the Association of Public Health Laboratories (APHL) for their technical assistance and vital role in the design and implementation of the wastewater-based surveillance activities. APHL teams supported laboratory staff training, protocol, and work plan development, and the coordination of data collection, analysis, and reporting activities.

## Abbreviations

CERSL: Central Emergency Response and Surveillance Laboratory.
CHAN: African Nations Championship
MG(s): Mass gathering(s).
NHLDS: National Health Laboratory and Diagnostic Services.
qPCR: Quantitative Polymerase Chain Reaction.
RCCE: Risk Communication and Community Engagement.
RSV: Respiratory Syncytial Virus.
WBS: Wastewater-based surveillance

## Declarations

### Ethics approval

This study was conducted under operational permission granted by the National Water and Sewerage Corporation, the agency responsible for Uganda’s water and sanitation infrastructure, to access wastewater collection points at Mandela National Stadium. Because wastewater-based surveillance does not involve collecting individually identifiable data from human participants, individual informed consent was not required. All wastewater sampling was conducted in accordance with Uganda’s Integrated Disease Surveillance and Response guidelines, and the study protocol was approved by the Uganda National Health Laboratory Services Research and Ethics Committee (Approval number: UNHLS-2025-133).

### Data availability

All data generated or analyzed from this study are included in this manuscript.

### Author contributions

Conceptualization: AN, SN

Methodology: AN, SN

Investigation: JM, CD, SS, RK

Data curation: AN, JK, IM

Validation: AN, JK, IM

Formal analysis: AN, JK, IM

Supervision: NH, VN, VU, DSA

Funding acquisition: FC, SN, NH

Resources: FC, SN, NH

Writing – original draft: AN, IM, VN, JK

Writing – review & editing: VB, HO, YKT, EB, SI, AM, HN, IS, CO, DA, JRA

All authors reviewed and approved the final manuscript.

